# Combined Utility of 25 Disease and Risk Factor Polygenic Risk Scores for Stratifying Risk of All-Cause Mortality

**DOI:** 10.1101/2020.03.13.20035527

**Authors:** Allison Meisner, Prosenjit Kundu, Yan Dora Zhang, Lauren V. Lan, Sungwon Kim, Disha Ghandwani, Parichoy Pal Choudhury, Sonja I. Berndt, Neal D. Freedman, Montserrat Garcia-Closas, Nilanjan Chatterjee

## Abstract

While genome-wide association studies have identified susceptibility variants for numerous traits, their combined utility for predicting broad measures of health, such as mortality, remains poorly understood. We used data from the UK Biobank to combine polygenic risk scores (PRS) for 13 diseases and 12 mortality risk factors into sex-specific composite PRS (cPRS). These cPRS were moderately associated with all-cause mortality in independent data: the estimated hazard ratios per standard deviation were 1.10 (95% confidence interval: 1.05, 1.16) and 1.15 (1.10, 1.19) for women and men, respectively. Differences in life expectancy between the top and bottom 5% of the cPRS were estimated to be 4.79 (1.76, 7.81) years and 6.75 (4.16, 9.35) years for women and men, respectively. These associations were substantially attenuated after adjusting for non-genetic mortality risk factors measured at study entry. The cPRS may be useful in counseling younger individuals at higher genetic risk of mortality on modification of non-genetic factors.

## INTRODUCTIONS

Genome-wide association studies (GWAS) with increasingly large sample sizes have led to the discovery of thousands of genetic variants associated with individual traits, including complex diseases and risk factors for disease (1). Analyses of polygenicity of a variety of traits (2,3) have further indicated that many individual traits are likely to be associated with thousands to tens of thousands of genetic variants, each with very small effect. Thus, much attention has been paid to the utility of polygenic risk scores (PRS), which represent the genetic burden of a given trait, for developing strategies for risk-based intervention through lifestyle modification (4-8), screening (5,7-12), and medication (5,7,13,14). A PRS for a given trait is typically defined as a weighted sum of a set of germline single-nucleotide polymorphisms (SNPs), where the weight for each SNP corresponds to an estimate of the strength of association between the SNP and the trait (7). Recent studies indicate that while PRS tend to have modest predictive capacity overall, they have the potential to offer substantial stratification of a population into distinct levels of risk for some common diseases such as coronary artery disease (CAD) and breast cancer (4,15).

There is ongoing debate regarding the utility of PRS in clinical practice (16-18). PRS can be more robust and cost-efficient tools for risk stratification than other biomarkers and risk factors. In particular, PRS do not change over time and thus need to be measured only once. Additionally, the risk associated with PRS for different traits appears in many cases to be fairly consistent over an individual’s life course (15,19) and time-varying lifestyle and clinical factors tend to act in a multiplicative way on baseline genetic risk (4,6,20,21). Further, if genome-wide genotype and/or sequencing data are available on an individual, the same data can be used to evaluate the PRS for a large number of traits simultaneously. Thus, beyond the use of PRS for prevention of specific diseases, it is important to evaluate their utility for broad health outcomes, particularly if PRS are to be utilized in routine health care.

The broad health impact of public health or clinical interventions is often measured in terms of their impact on all-cause mortality or lifespan (22-25). While a small number of genetic variants associated with lifespan have been identified (26-28), no study to date has systematically evaluated the ability of emerging PRS for life-threatening diseases and mortality risk factors to predict mortality. We used data from the UK Biobank, a large prospective cohort study, to assess the combined utility of PRS associated with 13 common diseases and 12 established risk factors for mortality. We used training data to combine the trait-specific PRS into sex-specific composite PRS (cPRS) that are predictive of all-cause mortality. We then evaluated the association of these cPRS with all-cause mortality and their ability to stratify mortality risk in independent test data. We also assessed the degree to which mortality risk associated with the cPRS was accounted for by mortality risk factors measured at the time of entry into the study, i.e., middle age for most participants. Finally, we examined the potential clinical use of the cPRS, namely, counseling individuals at higher genetic risk of mortality on modification of non-genetic risk factors such as body mass index (BMI) and smoking status.

## SUBJECTS AND METHODS

### Causes of Death and Mortality Risk Factors

We used the Centers for Disease Control (CDC) Wide-ranging ONline Data for Epidemiologic Research (WONDER) database to identify the top causes of death (organized by the International Classification of Diseases (ICD)-10 113 Causes List) in terms of the number of deaths among non-Hispanic whites in the United States over the age of 40 in 2017, separately for men and women (29). We then determined the top 10 causes of death with some genetic basis, i.e., causes for which there is evidence of an association between one or more genetic variants and disease risk (Table S1). These causes accounted for 70.3% and 71.8% of deaths among women and men, respectively, in the CDC data.

Several of these causes were very general categories of disease (e.g., “diseases of heart”), making it difficult to identify relevant trait-specific GWAS. Thus, we identified the specific cause within these categories associated with the highest number of deaths (with the exception of “malignant neoplasms”; here, we identified the top four cancers for each sex in terms of the number of deaths). The final list of diseases was: CAD, COPD, Alzheimer’s disease, stroke, type 2 diabetes, CKD, hypertension, alcoholic liver cirrhosis, Parkinson’s disease, pancreatic cancer, colorectal cancer, lung cancer, breast cancer (women only), and prostate cancer (men only) (Table S1). These causes of death captured 44.4% and 44.9% of deaths among women and men, respectively, in the CDC data. The difference between these figures and those cited above (70.3% and 71.8% for women and men, respectively) are driven largely by deaths from non-CAD diseases of the heart and deaths from malignant neoplasms not included in our list of cancers. As our analysis involves UK Biobank data, we also used Office of National Statistics mortality data (30) to determine the top causes of death in the UK; these were nearly identical to those identified using the CDC data (Table S1).

Based on government statistics from the UK (31), we further identified major mortality risk factors that are known to have some genetic component (32,33). We included smoking status, alcohol consumption, SBP, BMI, total cholesterol, fasting plasma glucose, and eGFR. Beyond the risk factors highlighted by the UK government statistics, we included LDL cholesterol, HDL cholesterol, triglycerides, DBP, and sleep duration. In particular, sleep duration was included on the basis of several studies showing clear links between sleep duration and all-cause mortality (34-36).

### Extraction of SNP Information from the GWAS Catalog and Publicly Available GWAS

To generate a PRS for each disease included in the top causes of death, we used results published in the NHGRI-EBI GWAS Catalog (37) to identify SNPs associated with the disease. We downloaded the GWAS Catalog results on March 15, 2019, and selected autosomal genome-wide significant SNPs (p-value ≤ 5 × 10^−8^). For each disease, we identified one or more search terms based on the trait names used by the GWAS Catalog, and selected the SNPs corresponding to these search terms. We then checked several fields of the GWAS Catalog, such as the source of the data, the study title, and the description of the trait studied, to ensure that we retained relevant SNPs; in particular, we sought to include results from analyses of Europeans (or multi-ethnic populations including Europeans) and to exclude studies of pleiotropic or composite outcomes, studies not of disease susceptibility, studies of children or pregnant women, studies of a secondary condition in individuals with a primary condition (e.g., myocardial infarction in individuals with coronary heart disease), studies of haplotypes or multi-SNP analyses, and studies of subpopulations (e.g., carriers of a specific genetic mutation; the only exceptions to this were studies of cirrhosis among alcohol drinkers and studies of COPD among smokers) or SNP-environment interactions. Importantly, these exclusions mean we included only GWAS of disease status, rather than GWAS of particular outcomes among individuals with a given disease, e.g., disease-associated mortality. In the resulting list of SNPs, there were several cases where the same SNP appeared multiple times for the same disease trait. In these situations, we kept the result from the largest study (in terms of the number of cases). The same SNP may appear for multiple traits.

For our analysis, it was important to extract the effect allele, effect size, and effect allele frequency for each SNP. The effect allele and effect size were used to construct the PRS in the UK Biobank, and the effect allele and effect allele frequency were used to check whether the SNP in the UK Biobank was the same as the SNP reported on the GWAS Catalog. For many SNPs on the list we created, some or all of this information was missing in the GWAS Catalog. We sought to fill in this information by consulting the original paper and its supplemental materials, as well as the Ensembl database (38). In situations where we were not able to discern the effect allele, the effect allele frequency, or the effect size of a particular SNP, the SNP was removed from our list.

We applied the same approach for identifying SNPs for each cause of death except for stroke. This is because there are several types of stroke and different studies included in the GWAS Catalog employed definitions of stroke with varying specificity. Thus, we used a recently published stroke PRS (39) instead of using the results available from the GWAS Catalog.

Our approach to identifying SNPs for inclusion in the mortality risk factor PRS differed from the approach described above. In particular, we found that the risk factor phenotypes were typically defined and/or analyzed differently across studies. For instance, smoking behavior could be defined as ever-use of cigarettes (never vs. former/current) or more granularly, incorporating cigarettes per day and duration among ever smokers. As another example, body mass index could be analyzed as a raw measurement, or it could first be rank-transformed. In light of these complications, instead of using the results included in the GWAS Catalog, we used the results from the most recent, largest trait-specific GWAS for which summary data were available (40-45). As above, we selected autosomal genome-wide significant SNPs (p ≤ 5 × 10^−8^) and removed SNPs for which the effect allele, effect size, or effect allele frequency were unavailable. In addition, as variant identifiers (RS IDs) were the primary way of querying the UK Biobank genotype data (described below), SNPs without RS IDs were removed (this was not an issue for the GWAS Catalog results).

### UK Biobank: Disease and Mortality Data

The UK Biobank is a large cohort study of over 500,000 individuals in the UK (46). The study enrolled individuals aged 40-69 years between 2006 and 2010 and has followed them since enrollment. A vast array of information has been collected from these individuals, including genotype data, anthropometric measurements, and information on lifestyle factors and personal and family history of disease. Additionally, data from national death and cancer registries are linked to the UK Biobank data.

We retrieved data on mortality, incident and prevalent disease for the top causes of death, and mortality risk factor measurements at baseline. The death registry data were available through November 30, 2016, for the centers in Scotland and January 31, 2018, for the centers in England and Wales. We determined whether an individual died of a particular disease by considering the ICD-10 code listed as the primary cause of death (see Table S1 for the codes used). We used several sources of data to identify incident and prevalent cases of disease for the top causes of death. In particular, we used cancer registry data (available through October 31, 2015, in Scotland and March 31, 2016, in England and Wales) to determine whether participants had or experienced the cancers in our list of diseases before (prevalent case) or after (incident case) study baseline on the basis of ICD-9 and ICD-10 codes (Table S2). For the non-cancer diseases, we used questionnaire/interview data, hospital episode data (available through March 31, 2017, in England, October 31, 2016, in Scotland, and February 29, 2016, in Wales), and death registry data to identify prevalent and incident cases of disease (Table S2). The exception to this was incident and prevalent diabetes, which were defined based on the algorithm presented in (47). For SBP and DBP at baseline, two measurements were made for each; when both of these were non-missing, the average was used. Self-reported intake of different forms of alcohol was converted into grams of alcohol per day (Table S3).

In all analyses, unless otherwise specified, we adjusted for the first ten genetic principal components, which were provided by the UK Biobank, in order to account for population stratification. In addition, all survival models accounted for left truncation by starting the follow-up interval at study entry. Throughout, we restricted our attention to unrelated participants (third degree relatives or closer were removed) of white British ancestry, in order to minimize the influence of population stratification and avoid issues related to clustering of individuals in families. We further removed individuals who had withdrawn their consent to participate. Unrelated participants were identified as those who were used by the UK Biobank to compute the principal components and ancestry was determined by the UK Biobank based on self-report and principal component analysis. The UK Biobank was approved by the North West Multi-centre Research Ethics Committee. This research was conducted using the UK Biobank Resource under Application Number 17712.

### Evaluating PRS in the UK Biobank

Imputed genotype data (in the form of allele dosage, i.e., between 0 and 2) for the SNPs identified above were extracted from the UK Biobank, matching on RS ID if possible and on chromosome and position otherwise. Non-biallelic SNPs and ambiguous palindromic SNPs (A/T or C/G SNPs with allele frequencies between 0.4 and 0.6) were removed. To ensure the SNPs from the UK Biobank were the same as those on our curated list of trait-associated SNPs, the alleles and allele frequencies were compared (allowing for the possibility of strand flips). SNPs that did not match the UK Biobank data, i.e., SNPs for which the reported allele frequency and the allele frequency in the UK Biobank differed by more than 0.15, were removed. Finally, SNPs in LD were removed via LD clumping, implemented using PLINK with an r^2^ cutoff of 0.1 and based on the reported p-values (from the GWAS Catalog or the publicly available summary statistics) and the 1000 Genomes European reference panel (48,49). This was done separately for each disease and risk factor, yielding a list of independent SNPs for each trait. The one exception was stroke: the SNP list was not pruned because the estimated association coefficients provided were based on a joint SNP model. The number of SNPs included in each PRS varied widely, between two SNPs for cirrhosis and 1,458 for BMI (Table S4). In total, our analysis included 3,941 unique SNPs.

Next, a PRS for each trait was constructed for each participant by weighting the SNP dosage by the reported log odds ratio (for binary traits) or linear regression coefficient (for continuous traits):

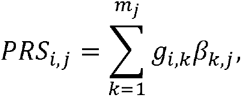

where *PRS_i,j_* is the PRS value for the i^th^ individual and the j^th^ trait, *m_j_* is the number of SNPs included in the PRS for the j^th^ trait, *g_i,k_* is the genotype dosage for the i^th^ individual and the k^th^ SNP, and *β_k,j_* is the log odds ratio or linear regression coefficient for the k^th^ SNP and the j^th^ trait.

### Statistical Analysis

All analyses were sex-specific and the PRS were standardized to have unit variance. We first evaluated the association between each derived PRS and the corresponding trait (i.e., prevalent disease and incident disease for the disease trait, and measurement at baseline for the mortality risk factors). For the disease traits, we evaluated the association with incident and prevalent disease status separately. To evaluate the relationship between each disease PRS and prevalent disease, we fit a logistic regression model for each disease. We used Poisson models with robust variance estimation (50) to evaluate the association between each disease PRS and incident disease among individuals without prevalent disease. For the mortality risk factors, we used linear regression with robust variance estimation to model the relationship between each mortality risk factor PRS and the risk factor measurement at baseline. The one exception was smoking status; since the smoking status PRS was developed based on a GWAS of ever-use of cigarettes, we defined the smoking status risk factor as ever-use of cigarettes. As this is a binary variable, we used logistic regression to model the relationship between the smoking status PRS and ever-use of cigarettes. Since eGFR was not directly available in the UK Biobank, we calculated eGFR at baseline using the Modification of Diet in Renal Disease (MDRD) Study equation (51); this mirrors the definition of eGFR used in the GWAS upon which our eGFR PRS was based (45). All models included adjustment for age at entry, in addition to the first ten principal components.

We also investigated cause-specific mortality for the diseases included in our top causes of death. We used Cox proportional hazards models to study the relationship between each disease PRS and age at death from that disease. Deaths from other causes were treated as censoring events. We performed these analyses in the full cohort and also among individuals with and without the disease corresponding to the cause of death being modeled at baseline.

We also evaluated the relationship between each mortality risk factor PRS and mortality due to each of the causes of death. For all of the analyses related to cause-specific mortality, when there were not enough deaths to yield stable estimates, estimates are not provided.

Our main analysis involved studying the joint relationship between the 25 PRS and all-cause mortality. First, we split the data into training (2/3) and test (1/3) sets. Then, in the training data, all PRS (with the exception of prostate cancer and breast cancer for the female- and male-specific models, respectively) were included in Cox proportional hazards models of age at death:

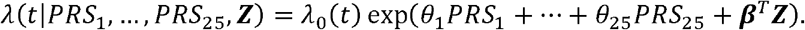

In this formula, *λ(t|PRS*_1_*,…,PRS*_25_) denotes the hazard at age *t* given *PRS*_1_*, …,PRS*_25_, λ_0_(*t*) denotes the baseline hazard at age *t*, and ***Z*** is a vector of the first ten principal components. Each model yielded a weighted combination of the individual PRS where the weights were the estimated log HRs from the Cox model, 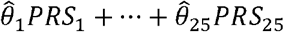; we refer to these sex-specific weighted combinations as the “composite PRS” (cPRS). These cPRS were then applied to the test data. In particular, we used a Cox model to evaluate the HR for all-cause mortality per standard deviation of the cPRS. In addition, we estimated the HR comparing individuals in the top 5% of the cPRS distribution to those in the middle 20% and the HR comparing individuals in the bottom 5% to those in the middle 20% in the test data. This was based on quantiles estimated in the training data. To aid in the interpretation of these results, the estimated HRs were converted into approximate years of life difference, as done in other studies of survival (26,33). In addition, we used Harrell’s C-index to quantify the discriminatory ability of the cPRS (52); note that this evaluation did not adjust for principal components.

We undertook a series of additional analyses. First, we evaluated the association between the cPRS and all-cause mortality in the “healthy” subset of the test data, that is, the test set after removing individuals with any of the diseases included as a top cause of death at baseline (i.e., prevalent cases). We also re-evaluated the association between the cPRS and all-cause mortality in the test data, adjusting for the mortality risk factors measured at baseline (that is, BMI, smoking status, alcohol consumption, SBP, DBP, eGFR, total cholesterol, LDL cholesterol, HDL cholesterol, triglycerides, blood glucose, and sleep duration), removing individuals in the test data that were missing any of these measurements. All risk factors were included as continuous variables, with the exception of smoking status, which was included as a binary variable (ever vs. never use).

Finally, we evaluated the relationship between two major modifiable risk factors, BMI and smoking status, and absolute risk of mortality for individuals at different levels of polygenic risk. We estimated the mortality risk for obese individuals (BMI > 30 kg/m^2^) and normal weight individuals (BMI of 18.5-25 kg/m^2^) based on Cox proportional hazards models with quintiles of the cPRS and BMI categories (≤ 18.5 kg/m^2^, (18.5-25 kg/m^2^], (25-30 km/m^2^], > 30 kg/m^2^), both modeled as categorical variables, fit in the test data. Estimates of risk for never smokers and ever smokers are based on Cox proportional hazards models with quintiles of the cPRS, modeled as a categorical variable, and an indicator of ever-use of cigarettes, fit in the test data. These models did not include adjustment for principal components.

All analyses were conducted using R (53), including the rms (54), survival (55), ggplot2 (56), and sandwich (57,58) packages. We report 95% confidence intervals throughout.

## RESULTS

### UK Biobank: Disease, Mortality, and Genotype Data

After removing individuals who were related, were not of British ancestry, or had withdrawn their consent to participate, our dataset included 337,138 participants, including 181,027 women and 156,111 men (Tables 1 and S5). There were 13,610 deaths (4.0%) with 5,250 among women (2.9%) and 8,360 among men (5.4%). The diseases included in the top causes of death accounted for 45.9% of the deaths in women and 45.5% of the deaths in men in the UK Biobank. Notably, very few deaths in the UK Biobank were attributed to type 2 diabetes, which appears to be due to many more deaths in the UK Biobank having type 2 diabetes listed as a secondary cause of death as opposed to the primary cause.

**Table 1:** Descriptive statistics. Descriptive statistics for the full cohort used for the analysis (after removing individuals who were related, were not of British ancestry, or had withdrawn their consent to participate), the training data (2/3 of the full cohort), and the test data (1/3 of the full cohort).

|  | Full cohort |  | Training data |  | Test data |  |
| --- | --- | --- | --- | --- | --- | --- |
|  | Women | Men | Women | Men | Women | Men |
| Sample size | 181,027 | 156,111 | 120,719 | 104,037 | 60,308 | 52,074 |
| Age at study entry (years; mean (SD)) | 57.2 (7.9) | 57.6 (8.1) | 57.2 (7.9) | 57.6 (8.1) | 57.2 (7.9) | 57.6 (8.1) |
| Follow-up (years; mean (SD)) | 8.8 (1.1) | 8.7 (1.3) | 8.8 (1.1) | 8.7 (1.3) | 8.8 (1.0) | 8.7 (1.3) |
| Number of deaths | 5,250 | 8,360 | 3,530 | 5,576 | 1,720 | 2,784 |

6 SD: standard deviation.

### Constructing and Evaluating the Trait-Specific PRS in the UK Biobank

As anticipated, the trait-specific PRS tended to be moderately to strongly associated with the corresponding disease or risk factor (Figure S1 and Table S6). The strongest associations for the disease traits (odds ratios or relative risks of at least 1.5 per standard deviation (SD)) were observed for Alzheimer’s disease (incident disease only), type 2 diabetes, breast cancer in women, prevalent CAD in men, cirrhosis in men, and prostate cancer in men.

We observed that the PRS for each disease was generally at least moderately associated with death from that disease (Figure 1), with the association being strongest for Alzheimer’s disease (hazard ratio (HR) per SD: 1.86 (95% confidence interval: 1.42, 2.42) in women; 2.01 (1.52, 2.65) in men), CAD (1.51 (1.34, 1.69) in women; 1.48 (1.40, 1.57) in men), breast cancer in women (1.51 (1.40, 1.63)), prostate cancer in men (1.68 (1.54, 1.84)), and cirrhosis in men (1.49 (1.03, 2.16)). In general, the PRS were stronger predictors of cause-specific mortality among individuals without prevalent disease than they were among individuals with prevalent disease (Figure S2); this indicates the PRS were typically more strongly associated with disease onset than with prognosis.

**Figure 1:**
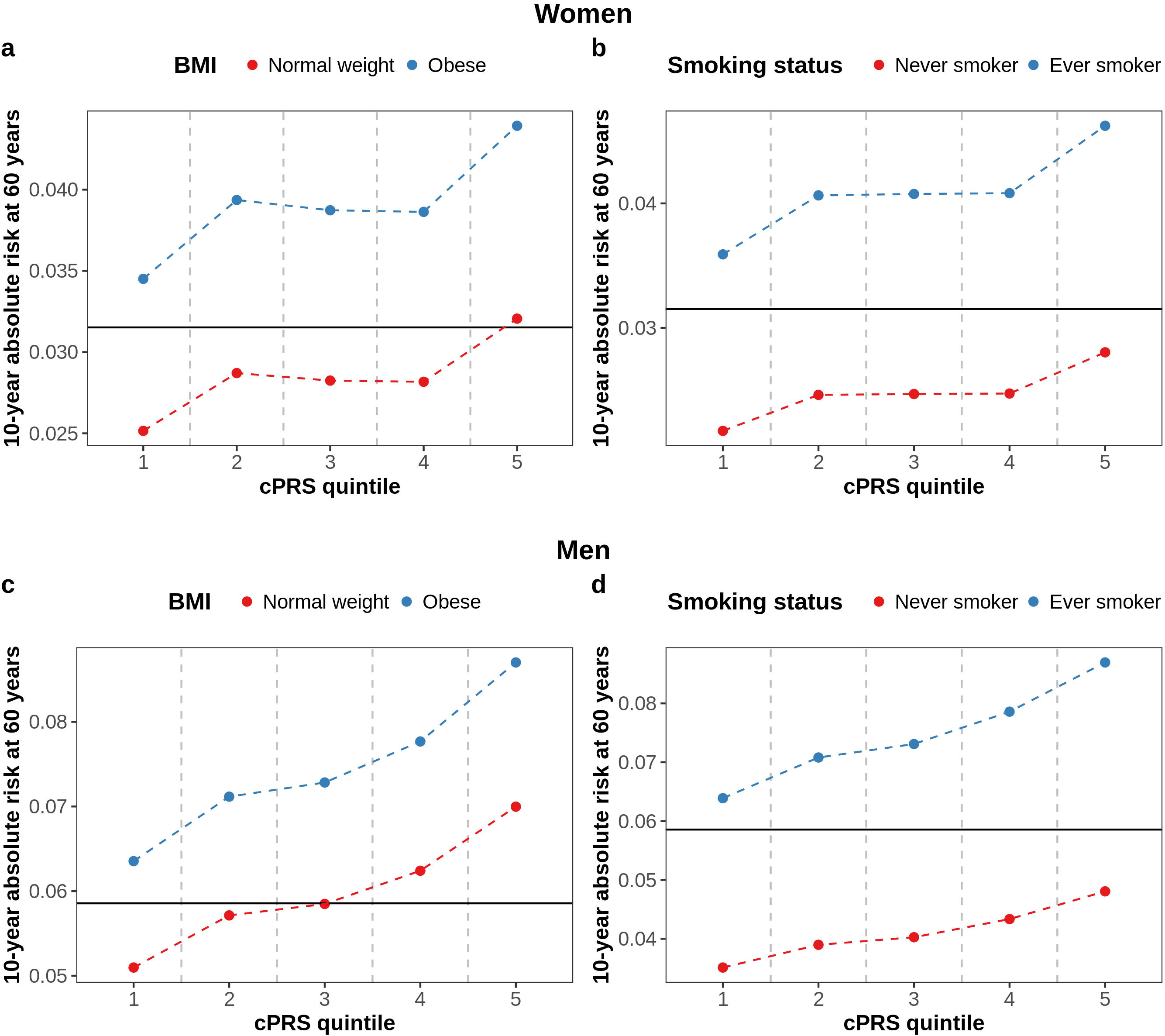
Association of each disease PRS with cause-specific mortality in the full cohort. For each disease, we evaluated the association between the disease PRS and mortality from the disease based on sex-specific Cox proportional hazards models of age at death. Deaths from other causes were treated as censoring events. Some causes did not have enough deaths to yield stable estimates, i.e., < 6 deaths; in these cases, estimates are not provided. Each PRS was standardized to have unit variance so the estimates correspond to the HR per SD of the PRS. The horizontal lines indicate 95% confidence intervals. CAD: coronary artery disease; COPD: chronic obstructive pulmonary disease; HR: hazard ratio; SD: standard deviation; PRS: polygenic risk score.

We found that the PRS for BMI was at least moderately associated with mortality related to CAD (primarily in men), COPD (among women), hypertension (among men), lung cancer (among women), pancreatic cancer (among women), Parkinson’s disease (among women), and stroke (among women) (Figures S3 and S4). The PRS for smoking was weakly associated with mortality due to CAD (among men) and moderately associated with mortality due to COPD (primarily in men) and lung cancer. The PRS for LDL cholesterol was strongly associated with mortality related to Alzheimer’s disease (among men) and COPD (among women) and moderately associated with mortality due to CAD (primarily in men). The PRS for total cholesterol was strongly positively associated with mortality due to Alzheimer’s disease (primarily in men) and COPD (among women), moderately positively associated with mortality related to CAD (among men), and moderately negatively associated with mortality due to pancreatic cancer (among men). The PRS for triglycerides was strongly negatively associated with mortality from stroke among men. The PRS for alcohol consumption was moderately positively associated with mortality due to CAD, primarily among men.

We found that several PRS were modestly associated with all-cause mortality, with some differences between men and women (Figure 2). The PRS for BMI was modestly associated with risk of all-cause mortality for both women (HR per SD: 1.07 (1.04, 1.10)) and men (1.08 (1.05, 1.10)). In addition, the PRS for smoking status, Alzheimer’s disease, LDL cholesterol, and lung cancer were modestly associated with all-cause mortality in both sexes. The PRS for breast cancer and prostate cancer were modestly associated with all-cause mortality in women and men, respectively. Among men, the PRS for CAD, cirrhosis, DBP, HDL cholesterol, SBP, stroke, total cholesterol, triglycerides, type 2 diabetes, and alcohol consumption were modestly associated with all-cause mortality; notably, the PRS for HDL cholesterol and triglycerides were both negatively associated with all-cause mortality. In general, the estimated associations tended to be stronger in men than in women.

**Figure 2:**
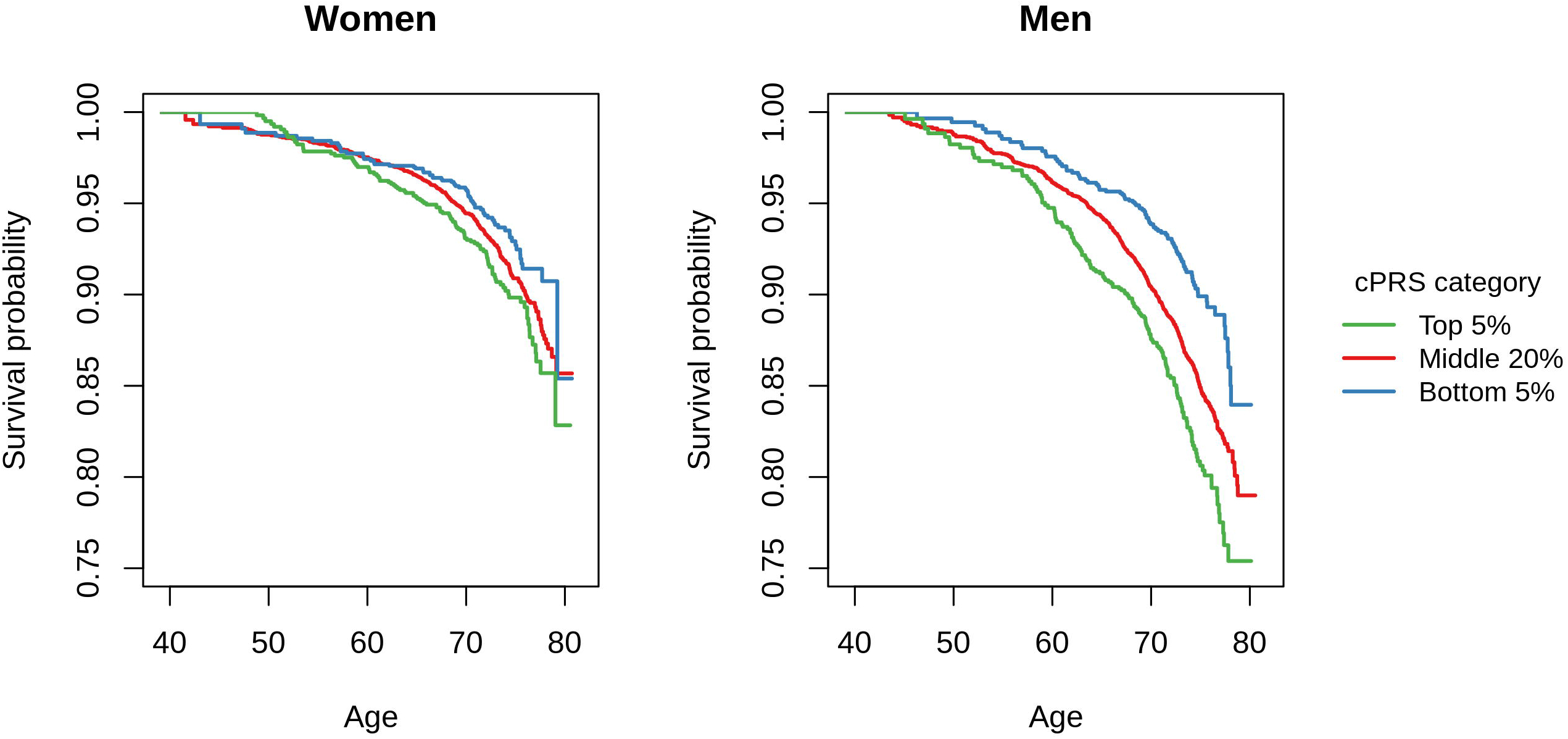
Association of each trait-specific PRS with all-cause mortality in the full cohort. We evaluated the association between each PRS and all-cause mortality based on sex-specific Cox proportional hazards models of age at death in the full cohort. Each Cox model included one PRS. Each PRS was standardized to have unit variance so the estimates correspond to the HR per SD of the PRS. The horizontal lines indicate 95% confidence intervals. BMI: body mass index; CAD: coronary artery disease; COPD: chronic obstructive pulmonary disease; DBP: diastolic blood pressure; eGFR: estimated glomerular filtration rate; HDL: high-density lipoprotein; LDL: low-density lipoprotein; SBP: systolic blood pressure; HR: hazard ratio; SD: standard deviation; PRS: polygenic risk score.

### Constructing and Evaluating the Composite PRS in the UK Biobank

The training data used the construct the cPRS included 224,756 participants, among them 120,719 women and 104,037 men (Table 1). There were 9,106 deaths in the training data with 3,530 in women and 5,576 in men. Correspondingly, the test data used to evaluate the cPRS included 112,382 individuals (60,308 women and 52,074 men) and 4,504 deaths (1,720 among women and 2,784 among men).

The cPRS were moderately associated with all-cause mortality in the test data (HR per SD: 1.10 (1.05, 1.16) in women, 1.15 (1.10, 1.19) in men; see Table 2 and Figure S5). However, the cPRS were able to identify substantial fractions of the population that have meaningfully elevated and reduced mortality risk, particularly among men (Table 2 and Figure 3). The estimated difference in life expectancy between the top and bottom 5% of the cPRS distribution was 4.79 (1.76, 7.81) years in women and 6.75 (4.16, 9.35) years in men. The overall discriminatory capacity of the cPRS, measured by Harrell’s C-index (52), was small: 0.525 in women and 0.536 in men. These are comparable to the values for several strong risk factors for mortality, including BMI (0.532 in women, 0.530 in men), smoking status (0.562 in women, 0.574 in men), and alcohol consumption (0.509 in women, 0.547 in men).

**Table 2:** The results of the main analysis of all-cause mortality and the cPRS, with and without adjustment for mortality risk factors. The cPRS were constructed in the training data and evaluated by fitting sex-specific Cox proportional hazards models of the association between the cPRS and age at death from all causes in the test data. Both the continuous cPRS and categorical cPRS were modeled. The estimated HRs and CIs were converted to estimated years of life lost. The analysis adjusting for mortality risk factors included adjustment for the risk factors measured at baseline (BMI, smoking status, alcohol consumption, SBP, DBP, eGFR, total cholesterol, LDL cholesterol, HDL cholesterol, triglycerides, blood glucose, and sleep duration); individuals missing any of these measurements were excluded.

|  | Women | Men |
| --- | --- | --- |
| **Without adjustment for mortality risk factors** |  |  |
| **Population in test data: N (deaths)** |  |  |
| Total population | 60,308 (1,720) | 52,074 (2,784) |
| Top 5% of cPRS | 3,060 (107) | 2,454 (159) |
| Middle 20% of cPRS | 12,005 (342) | 10,387 (539) |
| Bottom 5% of cPRS | 3,096 (69) | 2,526 (89) |
| **cPRS: HR (95% CI)** |  |  |
| Per SD of cPRS | 1.10 (1.05, 1.16) | 1.15 (1.10, 1.19) |
| Top 5% vs. middle 20% of cPRS | 1.24 (1.00, 1.54) | 1.27 (1.07, 1.52) |
| Bottom 5% vs. middle 20% of cPRS | 0.77 (0.59, 1.00) | 0.65 (0.52, 0.81) |
| Top 5% vs. bottom 5% of cPRS | 1.61 (1.19, 2.18) | 1.96 (1.52, 2.55) |
| **cPRS: years of life lost (95% CI)** |  |  |
| Per SD of cPRS | 0.97 (0.50, 1.44) | 1.36 (0.98, 1.73) |
| Top 5% vs. middle 20% of cPRS | 2.17 (0.00, 4.34) | 2.42 (0.65, 4.19) |
| Bottom 5% vs. middle 20% of cPRS | -2.61 (-5.20, -0.03) | -4.33 (-6.58, -2.09) |
| Top 5% vs. bottom 5% of cPRS | 4.79 (1.76, 7.81) | 6.75 (4.16, 9.35) |
| **With adjustment for mortality risk factors** |  |  |
| **Population in test data: N (deaths)** |  |  |
| Total population | 36,008 (855) | 36,283 (1,730) |
| Top 5% of cPRS | 1,799 (51) | 1,689 (102) |
| Middle 20% of cPRS | 7,143 (168) | 7,240 (329) |
| Bottom 5% of cPRS | 1,907 (37) | 1,804 (60) |
| **cPRS: HR (95% CI)** |  |  |
| Per SD of cPRS | 1.06 (0.99, 1.13) | 1.10 (1.04, 1.15) |
| Top 5% vs. middle 20% of cPRS | 1.19 (0.87, 1.63) | 1.25 (1.00, 1.56) |
| Bottom 5% vs. middle 20% of cPRS | 0.88 (0.62, 1.26) | 0.73 (0.55, 0.96) |
| Top 5% vs. bottom 5% of cPRS | 1.35 (0.88, 2.07) | 1.71 (1.24, 2.36) |
| **cPRS: years of life lost (95% CI)** |  |  |
| Per SD of cPRS | 0.58 (-0.11, 1.26) | 0.92 (0.43, 1.40) |
| Top 5% vs. middle 20% of cPRS | 1.72 (-1.43, 4.86) | 2.20 (-0.03, 4.43) |
| Bottom 5% vs. middle 20% of cPRS | -1.27 (-4.85, 2.30) | -3.19 (-5.95, -0.43) |
| Top 5% vs. bottom 5% of cPRS | 2.99 (-1.28, 7.26) | 5.39 (2.18, 8.60) |

BMI: body mass index; CI: confidence interval; cPRS: composite polygenic risk score; DBP: diastolic blood pressure; eGFR: estimated glomerular filtration rate; HDL: high-density lipoprotein; HR: hazard ratio; LDL: low-density lipoprotein; SBP: systolic blood pressure; SD: standard deviation.

**Figure 3:**
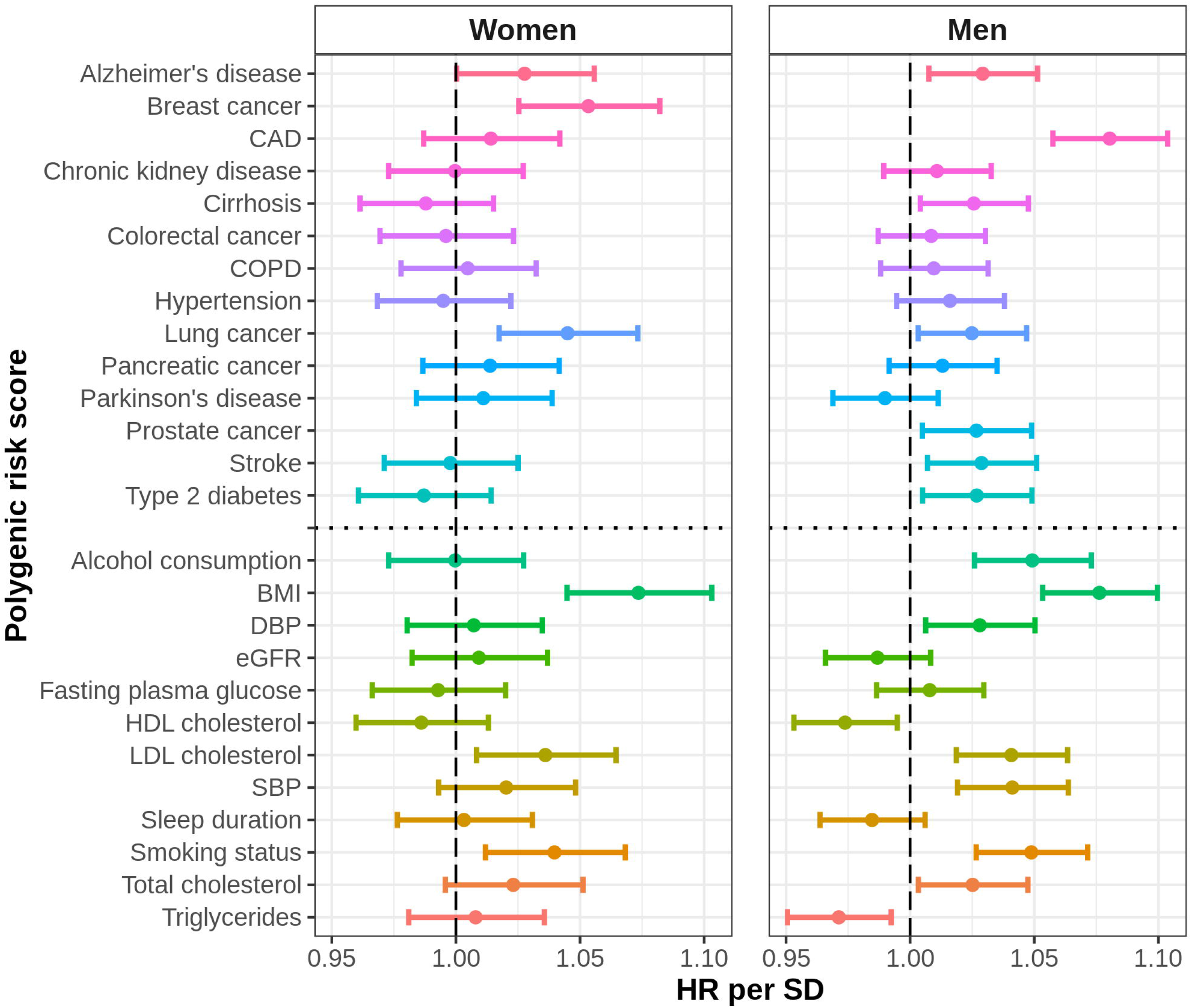
Kaplan-Meier survival curves by quantile of the cPRS. We estimated the sex-specific Kaplan-Meier survival curves for all-cause mortality by quantile of the cPRS in the test data. The Kaplan-Meier curves do not include adjustment for principal components. cPRS: composite polygenic risk score.

When we evaluated the cPRS in the “healthy” subset of the test data, the estimated associations between the cPRS and all-cause mortality were fairly similar to the results from the main analysis (Table S7). Separately, when we adjusted for the mortality risk factors measured at baseline, the association between the cPRS and all-cause mortality was markedly attenuated for both sexes (Table 2). These results indicate that a substantial fraction (40.7% for women and 32.5% for men) of the association between the cPRS and all-cause mortality was accounted for by these risk factors, which are (to varying degrees) heritable traits. After controlling for the measured risk factors, the difference in life expectancy between the top 5% and the bottom 5% of the cPRS distribution was estimated to be 2.99 (−1.28, 7.26) years in women and 5.39 (2.18, 8.60) years in men.

Finally, we evaluated the relationship between BMI and smoking status and absolute risk of mortality for individuals at different levels of polygenic risk (Figure 4). We observe that the estimated 10-year absolute risk of mortality for a 60-year-old woman in the top 20% of the cPRS distribution who is obese is 0.044. This is 38% higher than the estimated risk for a woman in the top 20% of the cPRS distribution who is not obese. Similarly, the estimated risk for a 60-year-old woman in the top 20% of the cPRS distribution who is a current or former is 64% higher than for a woman who has never smoked (0.046 vs. 0.028). Likewise, for a 60-year-old man, the estimated 10-year risk of mortality is 24% higher if the man is obese as opposed to normal weight (0.087 vs. 0.070) and the estimated risk is 81% higher if the man is a current or former smoker relative to a man who has never smoked (0.087 vs. 0.048). These differences highlight the potential importance of lifestyle modification even among those at high genetic risk. Furthermore, in most of these examples, the estimated risk for an individual who is in the top 20% of the cPRS distribution but who has a favorable risk factor profile is below the estimated risk for an individual in the middle 20% of the cPRS distribution, i.e., someone at moderate genetic risk (0.032 in women and 0.059 in men).

**Figure 4:**
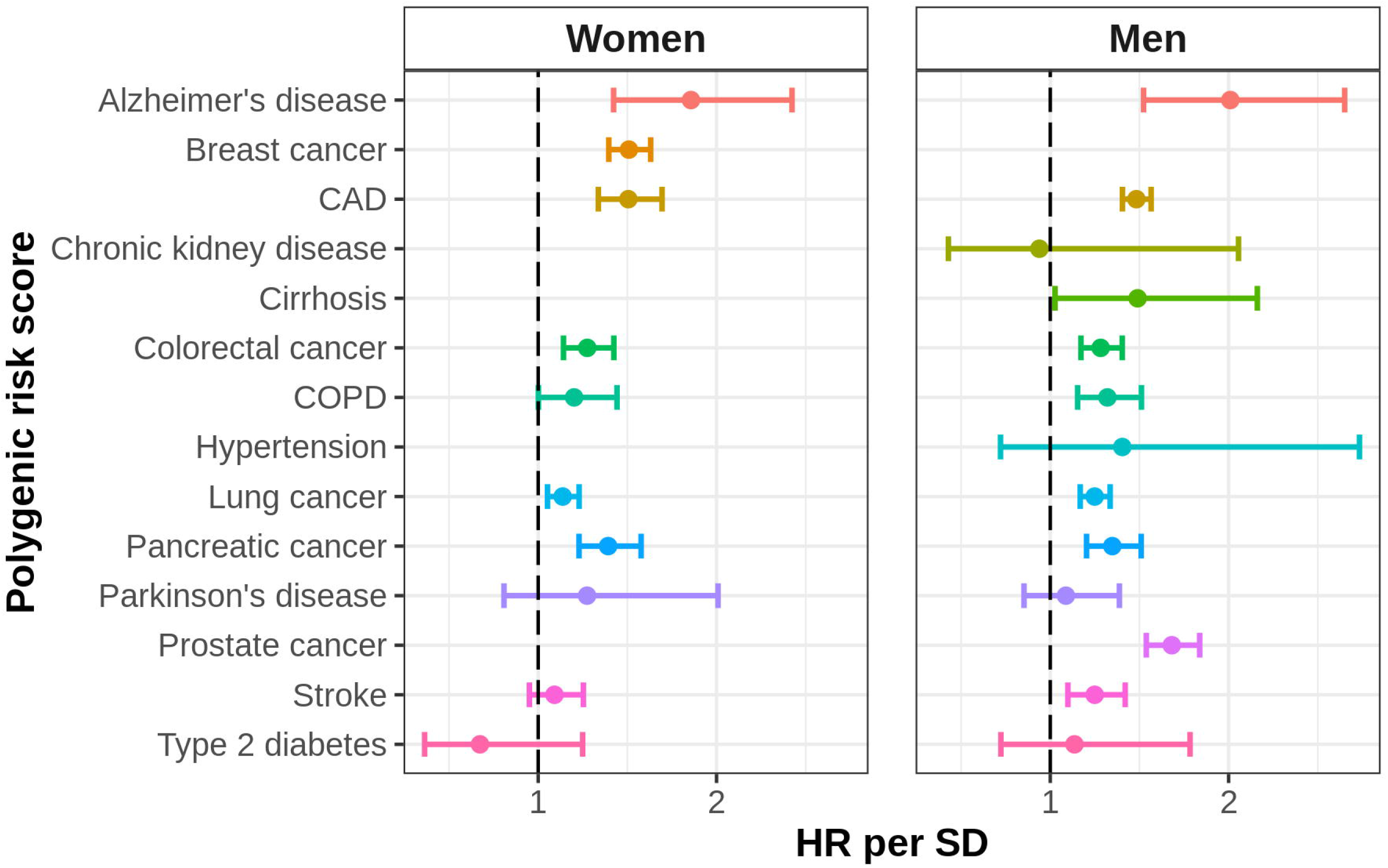
Estimates of absolute risk of mortality in different strata of the cPRS for specific categories of BMI and smoking status. We generated estimates of 10-year absolute risk of all-cause mortality for a 60-year-old in different strata of the cPRS for specific values of two mortality risk factors, BMI and smoking status. Panels A and B correspond to the results in women (for BMI and smoking status, respectively) and panels C and D correspond to the results in men (for BMI and smoking status, respectively). The horizontal line in each plot corresponds to an estimate of 10-year absolute risk of all-cause mortality for a 60-year-old in the middle quintile of the cPRS, based on sex-specific Cox proportional hazards models with quintiles of the cPRS, modeled as a categorical variable, fit in the test data. BMI: body mass index; cPRS: composite polygenic risk score.

## DISCUSSION

Analyses using a large dataset from the UK Biobank indicate that sex-specific composite PRS (cPRS) for all-cause mortality have fairly modest predictive capacity overall. However, there is evidence that the cPRS could identify substantial fractions of the population with notably elevated and reduced risk of all-cause mortality due to the genetic risk accumulated across many variants. Importantly, our results also show that a substantial proportion of the association between the cPRS and mortality was accounted for by mortality risk factors measured in middle age. These findings suggest that those individuals at high genetic risk of mortality may derive substantial benefit from modification of lifestyle factors; in particular, the cPRS could be useful in counseling individuals at high genetic risk on possible lifestyle choices that are associated with lower mortality risk.

A previous study evaluated the utility of 707 SNPs identified from GWAS of 125 diseases and risk factors for estimating mortality risk (32). This study developed a PRS directly from the individual SNPs, counting only the number of detrimental or protective alleles across the variants (i.e., without weighting the SNPs by the strength of association). In a combined analysis of men and women from two studies of northern European populations, the study reported a 10% higher risk of mortality between individuals in the 4^th^ versus 1^st^ quartile of the resulting PRS. In contrast, in the current study, we focus on a limited number of the most important causes of and risk factors for mortality, and build cPRS for mortality based on the underlying PRS. Our cPRS, although evaluated in a different population, appears to provide greater mortality risk stratification (HR for 4^th^ vs. 1^st^ quartile = 1.29 (1.13, 1.48) in women; 1.38 (1.24, 1.53) in men). These differences may be due to the incorporation of a larger number of SNPs emerging from more recent GWAS as well as the weighting of individual SNPs to account for their association with the individual diseases and risk factors in our analysis.

Several recent studies (26,59-62) have investigated the association of individual genetic variants and PRS with parental lifespans due to the increased power of these analyses relative to analyses of lifespan in genotyped individuals. Two large GWAS of parental lifespan, both including data from the UK Biobank, identified a total of only 18 loci (26-28), highlighting major challenges in finding individual variants related to lifespan. We constructed a lifespan PRS based on 17 of these variants (one was excluded as it was a palindromic SNP whose direction could not be resolved) and found modest associations with all-cause mortality (HR per SD: 1.02 (0.99, 1.05) in women and 1.04 (1.02, 1.06) in men). We further constructed a new cPRS, which included the 25 disease and risk factor PRS constructed for our analysis as well as the lifespan PRS; the associations of this new cPRS with all-cause mortality were nearly identical to that of the original cPRS (HR per SD of the new cPRS: 1.10 (1.05, 1.15) in women and 1.14 (1.10, 1.19) in men).

An important limitation of previous studies is the lack of adjustment for known mortality risk factors in characterizing the potential utility of PRS for estimating mortality risk. In our analysis, the association between the cPRS and mortality was attenuated by over 30% after adjusting for the mortality risk factors under study. These results suggest that while genetic variants associated with complex traits in GWAS could provide some mortality risk stratification early in life, their utility later in life, when other risk factors for mortality can be measured, is diminished.

Most GWAS are case-control studies of disease risk as opposed to prognosis, i.e., aggressiveness and/or progression of the disease leading to death. When we examined the association of the disease PRS with the corresponding cause-specific mortality among individuals with prevalent disease in the UK Biobank (Figure S2), only the PRS for CAD and COPD were (at least moderately) associated; in other words, for most PRS, there was little to no evidence of an association with prognosis or disease survival. Although such analyses may be influenced by selection associated with survivorship and poor health, in general, there is little evidence of association between disease risk SNPs (and thus disease PRS) and survival following disease onset. While future GWAS focusing on genetic determinants of aggressiveness and disease progression are needed, finding associations may be challenging due to available sample sizes and heterogeneity as a result of various factors such as treatment.

Our analysis of the relationship between the individual PRS and all-cause mortality revealed some important patterns (Figure 2). The strongest positive associations were seen for the PRS for BMI, breast cancer (in women), CAD (in men), smoking status (particularly in men), and alcohol consumption (in men). In addition, weaker associations with all-cause mortality were seen for the PRS for Alzheimer’s disease, lung cancer, and LDL cholesterol in both sexes and, among men, associations were seen for the PRS for stroke, cirrhosis, total and HDL cholesterol, prostate cancer, triglycerides, SBP, DBP, and type 2 diabetes. The negative association observed among men for the triglycerides PRS appears to be driven by a strong negative association between the triglycerides PRS and stroke-specific mortality (Figure S4), which is consistent with the “triglycerides paradox” reported by others (63-66).

Given that the associations of the CAD PRS with CAD-specific mortality were similar for men and women, the differences in the associations with all-cause mortality may be due to lower rates of CAD in women during the relatively short follow-up period of the UK Biobank. Differential event rates for some diseases for which alcohol consumption is a risk factor (e.g., CAD) could also partially explain the differences observed in the association of the alcohol consumption PRS with all-cause mortality by sex. We note that the sex differences observed in our results more generally are supported by other studies, which have similarly found indications of differences between men and women in the mechanisms governing lifespan and longevity (26,27,33,60,61,67,68).

Our results are generally consistent with a recent paper looking at PRS for many clinical risk factors and mortality across the UK Biobank, a Finnish biobank (FinnGen), and Biobank Japan (69). In this multi-ethnic study, several modest associations were observed, including for the PRS for SBP, DBP, and BMI (HRs of around 1.03-1.04 per SD in the trans-ethnic meta-analysis). Interestingly, the results from this analysis varied by ethnicity: for instance, within the UK Biobank, the association between the PRS for BMI and mortality reported in Sakaue et al. (69) was stronger than was observed in the trans-ethnic meta-analysis (HR of approximately 1.07 per SD in the UK Biobank versus 1.04 in the meta-analysis). This highlights the importance of multi-ethnic analyses.

We evaluated the broad utility of PRS in terms of their combined ability to predict mortality. In the future, other broad measures of health outcomes and expenditures, such as disability-adjusted life years (DALYs), should also be considered. The framework we have created for combining individual PRS could be used to a create composite PRS for DALYs or other measures. Given that PRS are known to be strongly associated with incidence of many debilitating diseases, one would anticipate such a composite PRS will have greater utility for predicting DALYs than for mortality. However, analysis of DALYs in a cohort study with limited follow-up, like the UK Biobank, is challenging.

Our analysis has several strengths. We used data from the UK Biobank, a large cohort study, to carry out a comprehensive analysis of PRS for complex traits and mortality, both overall and cause-specific. We used a novel approach to derive composite PRS across many diseases and risk factors to evaluate their combined utility for predicting overall mortality. Under the assumption that common genetic variants identified through recent GWAS influence mortality risk through the outcomes underlying the GWAS, the composite PRS approach provides a more parsimonious and powerful approach to building models for predicting composite outcomes than building models based on individual SNPs. The weights of individual SNPs in a PRS account for the strength and direction of association of each SNP with the corresponding outcome and the weights for the individual PRS in the cPRS reflect (in part) the relative contribution of the individual diseases and risk factors to mortality. Further, we conducted an unbiased evaluation of the performance of the cPRS for predicting mortality by building it in a training dataset and evaluating it in an independent test dataset.

As the UK Biobank participants are volunteers, there is evidence that this cohort differs from the general UK population in important ways, including being less likely to be obese, smoke, or drink alcohol (70). Selection bias (70), which contributes to such differences, could influence the generalizability of our results (71). Additionally, while our cPRS include germline mutations and so could potentially be evaluated at birth, the UK Biobank is comprised of individuals who have survived to at least middle age. Consequently, the results may not be fully generalizable to younger individuals and must be validated in other populations. Furthermore, the analysis of the cPRS with adjustment for the mortality risk factors required excluding observations in the test data with missing values for any of these risk factors. These observations constituted a substantial portion of the test data (40.3% in women, 30.3% in men). However, as the missingness mechanism for at least some risk factors is expected to be not random (e.g., individuals choosing not to answer questions regarding smoking status or alcohol consumption due to the social stigma surrounding these behaviors), imputation is not appropriate. Thus, some caution is warranted in interpreting these results.

As our analysis involved the evaluation of a large number of associations, issues related to multiple comparisons are a potential concern. However, our main analysis of the cPRS was carefully defined a priori and performed in independent test data. The other analyses we performed were intended to check the validity of the PRS we developed and to better understand the results of the main analysis of the cPRS. Additionally, we emphasize the strength of association rather than statistical significance in interpreting the results throughout. Another potential limitation of this analysis was our use of the GWAS Catalog to identify SNPs for inclusion in the disease PRS. As the GWAS Catalog is not an exhaustive listing of SNPs associated with every trait, we may have missed some associated SNPs. However, we believe that our approach, which allowed us to apply a uniform procedure for SNP selection to all diseases, captured most of the genetic susceptibility for each disease, and any differences in the PRS would be minor. Even if our PRS included all susceptibility SNPs identified by GWAS, the ability of the trait-specific PRS to predict all-cause mortality is related to both the power of the GWAS as well as the genetic correlation between the trait studied in the GWAS and all-cause mortality (72). Consequently, as GWAS continue to increase in power, we may find that trait-specific PRS are more strongly associated with all-cause mortality. In addition, further research on the genetic determinants of disease prognosis and survival may increase the utility of PRS in understanding mortality risk.

In conclusion, our results suggest that by combining knowledge gained from GWAS of complex traits, it may be possible to identify individuals who are expected to live substantially longer or shorter. In light of the ethical repercussions of using genetics to make predictions regarding an individual’s life course at birth, we argue that the cPRS may be most useful for counselling individuals about their genetic risk. In particular, the results of our analysis highlight the importance of considering genetic risk in the context of clinical risk factors measured in adulthood; thus, the cPRS may be useful in advising patients on the importance of certain lifestyle choices associated with mortality risk. Using the cPRS in this way would require validation of the cPRS outside of the UK Biobank.

## Data Availability

All data used are available from the UK Biobank upon request.

https://www.ukbiobank.ac.uk

## SUPPLEMENTAL DATA DESCRIPTION

The Supplemental Data includes seven tables and five figures.

## ACKNOWLEDGMENTS

This research has been conducted using the UK Biobank Resource under Application Number 17712. This research was supported by the Patient-Centered Outcomes Research Institute (ME-1602-34530); the National Institutes of Health (1 R01 HG010480-01); and the Intramural Research Program, Division of Cancer Epidemiology and Genetics, National Cancer Institute.

